# Exposome-wide association study of cognition among older adults in the National Health and Nutrition Examination Survey

**DOI:** 10.1101/2024.07.19.24310725

**Authors:** Lauren Y. M. Middleton, Erika Walker, Scarlet Cockell, John Dou, Vy K. Nguyen, Mitchell Schrank, Chirag J. Patel, Erin B. Ware, Justin A. Colacino, Sung Kyun Park, Kelly M. Bakulski

## Abstract

Cognitive impairment among older adults is a growing public health challenge and environmental chemicals may be modifiable risk factors. A wide array of chemicals has not yet been tested for association with cognition in an environment-wide association framework. In the US National Health and Nutrition Examination Survey (NHANES) 1999-2000 and 2011-2014 cross-sectional cycles, cognition was assessed using the Digit Symbol Substitution Test (DSST, scores 0-117) among participants aged 60 years and older. Concentrations of environmental chemicals measured in blood or urine were log_2_ transformed and standardized. Chemicals with at least 50% of measures above the lower limit of detection were included (n_chemicals_=147, n_classes_=14). We tested for associations between chemical concentrations and cognition using parallel survey-weighted multivariable linear regression models adjusted for age, sex, race/ethnicity, education, smoking status, fish consumption, cycle year, urinary creatinine, and cotinine. Participants with at least one chemical measurement (n=4,982) were mean age 69.8 years, 55.0% female, 78.2% non-Hispanic White, and 77.0% at least high school educated. The mean DSST score was 50.4 (standard deviation (SD)=17.4). In adjusted analyses, 5 of 147 exposures were associated with DSST at p-value<0.01. Notably, a SD increase in log_2_-scaled cotinine concentration was associated with 2.71 points lower DSST score (95% CI -3.69, -1.73). A SD increase in log_2_-scaled urinary tungsten concentration was associated with 1.34 points lower DSST score (95% CI -2.11, -0.56). Exposure to environmental chemicals, particularly heavy metals and tobacco smoke, may be modifiable factors for cognition among older adults.

## Introduction

Mild cognitive impairment is characterized by greater cognitive decline than expected based on age and educational attainment.^1^ In the United States, women experience a 71% lifetime risk of developing cognitive impairment at an average age at incidence of 73.^2^ Similarly, men experience 61% lifetime risk of cognitive impairment at an average age at incidence of 70.^2^ When these cognitive impairments impact activities of daily living, their syndrome has progressed to dementia, and a common subtype of dementia is Alzheimer’s disease.^3,4^ These are prevalent and challenging conditions, and understanding modifiable risk factors is key to public health prevention.

Later life environmental exposures such as air pollution and cigarette smoking are key modifiable factors for dementia prevention highlighted by the 2020 Lancet Commission.^5^ Additional environmental factors are suggestive based on current evidence. For example, among 2,023 older adults in the United States, a 0.51ng/mL increase in urinary cadmium concentrations was associated with 1.58 times higher hazard of Alzheimer’s disease mortality (95% CI: 1.20, 2.09) over 7.5 years of follow up.^6^ In addition, among 741 older men in the Boston area, a 21μg/g increase in patella bone lead was associated with -0.016 units of loss on the Mini Mental Status Exam score per year (95% CI: -0.032, -0.0004) over 15 years.^7^ Environmental chemical exposures are prevalent and potentially neurotoxic, though prior studies of environmental associations with cognition have been limited to few individual chemicals.

In genomic research, examining “well lit” areas of the genome based on prior evidence is described as the streetlight effect, and which may cause investigators to miss important but currently “dimmer” areas.^8^ Similarly, in environmental chemical research it is important to balance hypothesis testing of well-known chemicals with discovery analyses to identify novel chemical associations. The exposome-wide association study (ExWAS) approach allows the simultaneous investigation of many chemicals in parallel models.^9,10^ This study sought to build on evidence on individual chemicals and cognitive impairment by using ExWAS to examine a wide variety of chemicals and chemical classes for association with cognitive status. We used a cross-sectional and nationally representative sample of older adults from the United States National Health and Nutrition Examination Survey (NHANES). Findings from this study may help generate new hypotheses for mechanistic, validation, and prevention research.

## Methods

### Study Design and Participants

This analysis is a cross-sectional exposome-wide association study (ExWAS) of data pooled across three cycles of NHANES. Since 1999-2000, NHANES is conducted biannually by the National Center for Health Statistics (NCHS) and is nationally representative of non-institutionalized civilians in the United States.^11^ Data are collected through questionnaires, clinical measurements, and biomarker measurements, including chemical concentrations. Every two years, data on independent samples of participants are released in cycles. Cognition in adults aged 60 years and older was assessed in four cycles: 1999-2000, 2001-2002, 2011-2012, and 2013-2014. We included three of these cycles (1999-2000, 2011-2012, and 2013-2014; N = 29,896). We excluded the 2001-2002 cycle because a key covariate, fish consumption, was not measured in older adults in that cycle. Raw data are publicly available through the NCHS (https://www.cdc.gov/nchs/nhanes/index.htm). Processed data are publicly available through Kaggle, figshare, and Hugging Face repositories.^12–15^ Participants provided informed consent at the time of participation in NHANES. The University of Michigan Institutional Review Board (IRB) approved these secondary data analyses (HUM00116291).

### Chemical Biomarker Measures

NHANES participants provided urine and blood samples in the Mobile Examination Centers. These samples were analyzed for a variety of biomarkers used to indicate exposures. Information on the laboratory methods used for measuring chemical biomarker concentrations in each cycle year are available through the NCHS.^16^ Across the three cycles of interest, 395 environmental biomarkers were available for analysis. The environmental biomarker data were quality controlled similar to our previously described process.^17,18^ Briefly, environmental biomarkers were first excluded if they were categorized as dietary components or phytoestrogens, or if they were measured only in smokers. Urinary cadmium measures that had interference with urinary moldybdenum were removed.^19,20^ Next, for any lipid-soluble biomarkers measured in blood, we used the lipid-adjusted measurements and excluded the unadjusted versions. Lastly, we excluded biomarkers if more than 50% of measures were below the reported lower limit of detection (LOD). All measurements below the chemical-specific LOD were imputed as LOD/√2 by NHANES. In total, 147 chemical biomarkers were included as exposures in the analysis. The chemical biomarkers were not all measured in the same cycles nor in the same participants. Consistent with prior analyses, chemical measures were log_2_ transformed for analysis.^18^ After transformation, the measures were also standardized (mean = 0, standard deviation = 1) within each chemical to enable comparisons between chemicals with different units and distributions.

### Cognitive Measures

Cognitive testing via the Digit Symbol Substitution Test (DSST) was performed among participants aged 60 years and older.^21^ The DSST consists of a set of symbols corresponding to the numbers 1 through 9. Participants are given a paper with rows of numbers and are asked to draw the matching symbols beneath each number. The test is scored based on the number of symbols that were drawn correctly in the allotted 120 seconds. In NHANES, the maximum possible score was 133 points.^22^ Our primary analysis used DSST as a continuous measure. As a sensitivity analysis, consistent with previous studies, we categorized DSST score of less than or equal to the 25^th^ percentile (score ≤ 29 in our sample) as mild cognitive impairment (MCI) and DSST greater than the 25^th^ percentile as normal cognition.^23^

### Covariate Measures

Covariates were collected by interview including age (years), sex (male, female), race/ethnicity (Mexican American, Other Hispanic, Non-Hispanic White, Non-Hispanic Black, Other Race/Multiracial), education (less than 9^th^ grade, 9-12^th^ grade without diploma, high school graduate/General Education Development degree, some college or Associate of Arts degree, college graduate or above), history of tobacco use (never, ever smoked at least 100 cigarettes, current use), history of alcohol consumption (consumption in the past 12 months), fish and shellfish consumption (number of times eaten in the past 30 days), and the cycle year of participation. To protect participant anonymity, NHANES top-coded ages greater than 85 years in cycles prior to 2007 as age 85 years. Those who reported an age greater than 80 years in 2007 or later were top-coded as age 80 years. Education was recategorized as a binary variable: less than high school and high school/GED or above. Never smokers were categorized as smoking fewer than 100 cigarettes, former smokers were categorized as smoking over 100 cigarettes but not currently, and current smokers were categorized as reporting current use. The units for alcohol consumption were scaled to the average number of drinks per month based on the past 12 months. The number of fish and shellfish eaten over 30 days were combined into one value and categorized into three groups: 0, 1-3, and 4+. We considered the following categories as reference groups: male, Non-Hispanic White, less than high school, no fish consumption, never smoker, and the first survey cycle with measurements for each chemical.

Waist circumference (cm) was measured in the Mobile Examination Centers by trained study staff. Urinary creatinine concentrations (mg/dL), serum creatinine concentrations (mg/dL), and serum cotinine concentrations (ng/mL) of the participants were also measured from provided samples. Similar to the exposure measures, urinary creatinine and serum cotinine were log_2_ transformed and standardized. For chemicals measured in urine, we used urinary creatinine as a covariate to account for urine dilution.^24^ Cotinine was included as a covariate for all chemicals other than smoking-related chemicals (e.g. hydroxycotinine and total cotinine) as a biomarker of cigarette smoke exposure. Estimated glomerular filtration rate (eGFR) was calculated from serum creatinine measurements using the Chronic Kidney Disease Epidemiology Collaboration (CKD-EPI) equation, excluding the term for race adjustment.^12,25,26^ We categorized normal kidney function as eGFR ≥60 ml/min/1.73 m^2^.^27^

### Statistical Analysis

All analyses were performed in R version 4.3.0 and code files are available (https://github.com/bakulskilab/Cognition_ExWAS).^28^ To account for the NHANES complex sampling design, we applied survey weights to all analyses using the R package ‘survey’, unless otherwise noted.^29^

Participants were excluded from cognitive testing by NHANES if they were aged less than 60 years at the time of assessment or by our study team if they had no chemical biomarker measures. Participant and chemical biomarker inclusion and exclusion were visualized using flow charts. The distributions of continuous covariates were described by mean and standard deviation, while categorical covariates were described by proportions. We compared distributions of descriptive characteristics for the included and excluded participants. Within the included analytic sample, we compared the distributions of covariates by NHANES cycle and by cognitive impairment status.

We calculated summary statistics for each chemical biomarker (participant count, minimum and maximum concentrations, weighted arithmetic and geometric means and standard deviations, weighted median, weighted 25^th^ and 75^th^ percentiles, and percentage of measurements above the LOD). We used a scatterplot to compare the sample size per chemical versus the percentage of measurements above the LOD, colored by chemical family.

We imputed missing covariate and outcome data using multiple imputation by chained equations (MICE) with the R package ‘mice’, creating m = 5 complete datasets.^30^ Due to the variation in sample sizes for the chemical exposure measures, we did not impute exposure data. Additionally, we did not impute eGFR to maintain a consistent division between normal and abnormal kidney function. Characteristics of the imputed variables are summarized in a descriptive table (**Supplemental Table 1**).

To test the associations between chemical exposures and continuous DSST scores, we used survey-weighted linear regression models adjusted for age (continuous), sex (categorical), race/ethnicity (categorical), education level (categorical), urinary creatinine (continuous), serum cotinine (continuous), reported smoking status (categorical), seafood consumption (categorical), and survey cycle (categorical).

Consistent with an ExWAS design, each chemical association was analyzed individually in a separate regression model.^10^ Of note, because biomarkers were measured in different subsets of the analytic sample, these regression models had varying sample sizes. Urinary creatinine was included only in models for urinary chemical measurements. Models for smoking-related compounds (serum cotinine, total cotinine, hydroxycotinine, and thiocyanate) were not adjusted for cotinine. To account for the MICE procedure, models were run separately on each of the five unique imputed datasets and pooled to produce one set of coefficient estimates per exposure.^31^ The chemical biomarker regression coefficients, 95% confidence intervals (CI), and p-values for each exposure were compiled into a table. Because the chemical concentrations were log_2_ transformed and standardized prior to modeling, the regression coefficients are interpreted as the adjusted association with DSST per standard deviation of the log_2_-scaled chemical concentration. For discovery analyses, we prioritized chemicals with an unadjusted p-value < 0.01. To account for multiple comparisons, we controlled the false discovery rate (FDR) using the Benjamini-Hochberg method with the ‘p.adjust’ function in the stats R package, and used a significance threshold of FDR < 0.05.^28,32^ We visualized the regression coefficients and statistical significance for all chemicals using a volcano plot and a forest plot. For the exposure associations with DSST, the coefficient estimates and confidence intervals for all are presented in tables.

### Sensitivity analyses

We developed four sensitivity analyses to further explore the associations between chemical biomarkers and DSST. First, kidney function is associated with both chemical exposures and cognitive function.^33–35^ We subset the sample to participants with normal kidney function as indicated by eGFR ≥60 ml/min/1.73 m^2^ and re-ran the models for each chemical. Second, to account for possible confounding by lifestyle characteristics, in the full sample, we further adjusted models for waist circumference (continuous) and alcohol consumption (categorical). Third, to present a more clinically relevant analysis, we used MCI status categories as a binary outcome variable. We conducted modified Poisson regression for each chemical, adjusted for the same covariates as the main analysis and reported the relative risks (RR) and 95% CI. Lastly, we again restricted to participants with normal kidney function and re-ran the Poisson regression models. The results for all sensitivity analyses were visualized in volcano plots and the regression output was compiled in tables.

## Results

### Participant Characteristics and Chemical Descriptions

The analysis included N = 4,982 participants across three NHANES cycles (**Figure 1A**). Within the included sample, the mean age was 69.8 years, 55% were female, and 78.2% were Non-Hispanic White (**Table 1**). On average, participants from cycle year 1999-2000 had lower observed DSST scores compared to participants from cycle years 2011-2014 (**Table 1**). The included participants were on average more likely to be Non-Hispanic White and had a higher DSST score compared to the excluded participants aged 60 years and older (**Supplemental Table 2**). Participants with MCI were older, less likely to be Non-Hispanic White, and less likely to have completed high school compared to participants without impairment (**Supplemental Table 3**).

**Figure 1.**
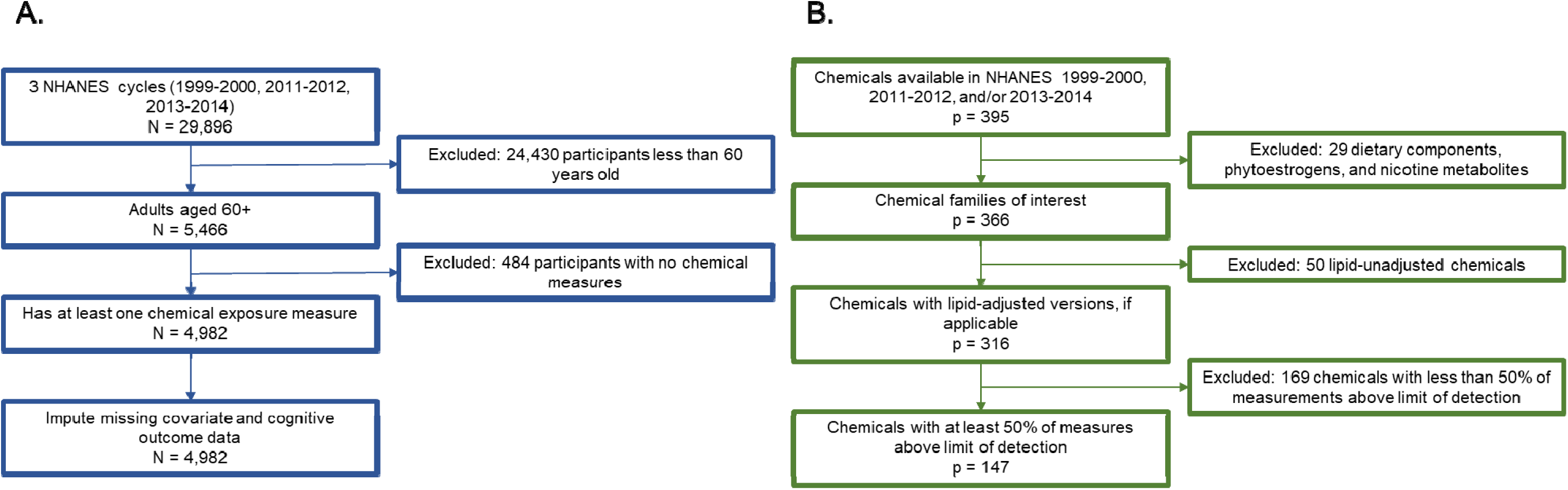
Inclusion criteria for participants (**A.**) and chemical biomarker measures (**B.**) in the National Health and Nutrition Examination Survey (NHANES). The Digit Symbol Substitution Test (DSST) was administered to participants aged 60+ years in the cycles 1999-2000, 2001-2002, 2011-2012, and 2013-2014. Cycle 2001-2002 was not considered because a key covariate (fish and seafood consumption) was not measured in the population of interest.

**Table 1.**
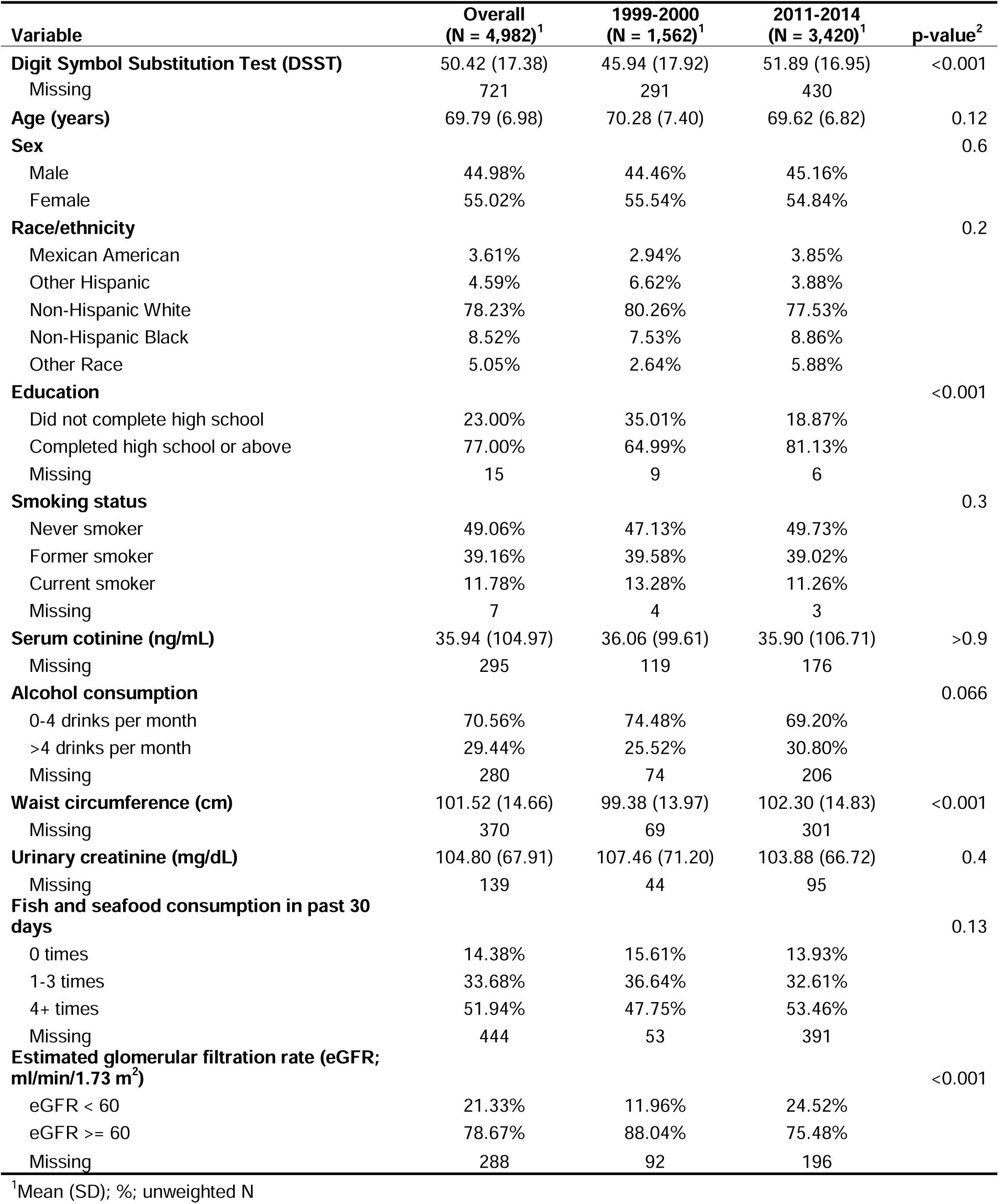
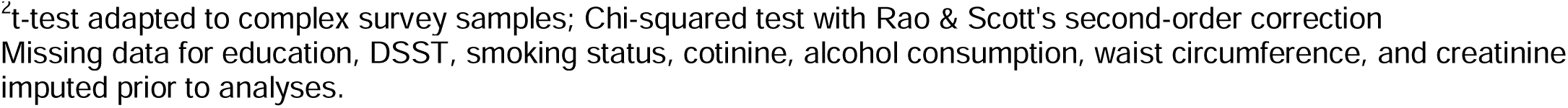
Weighted descriptive statistics of included participants in the National Health and Nutrition Examination Survey (NHANES), overall and stratified by earlier (1999-2000) versus later (2011-2014) cycles of participation (N = 4,982).

| Variable | Overall<br>(N = 4,982)<sup>1</sup> | 1999-2000<br>(N = 1,562)<sup>1</sup> | 2011-2014<br>(N = 3,420)<sup>1</sup> | p-value<sup>2</sup> |
| --- | --- | --- | --- | --- |
| **Digit Symbol Substitution Test (DSST)** | 50.42 (17.38) | 45.94 (17.92) | 51.89 (16.95) | <0.001 |
| Missing | 721 | 291 | 430 |  |
| **Age (years)** | 69.79 (6.98) | 70.28 (7.40) | 69.62 (6.82) | 0.12 |
| **Sex** |  |  |  | 0.6 |
| Male | 44.98% | 44.46% | 45.16% |  |
| Female | 55.02% | 55.54% | 54.84% |  |
| **Race/ethnicity** |  |  |  | 0.2 |
| Mexican American | 3.61% | 2.94% | 3.85% |  |
| Other Hispanic | 4.59% | 6.62% | 3.88% |  |
| Non-Hispanic White | 78.23% | 80.26% | 77.53% |  |
| Non-Hispanic Black | 8.52% | 7.53% | 8.86% |  |
| Other Race | 5.05% | 2.64% | 5.88% |  |
| **Education** |  |  |  | <0.001 |
| Did not complete high school | 23.00% | 35.01% | 18.87% |  |
| Completed high school or above | 77.00% | 64.99% | 81.13% |  |
| Missing | 15 | 9 | 6 |  |
| **Smoking status** |  |  |  | 0.3 |
| Never smoker | 49.06% | 47.13% | 49.73% |  |
| Former smoker | 39.16% | 39.58% | 39.02% |  |
| Current smoker | 11.78% | 13.28% | 11.26% |  |
| Missing | 7 | 4 | 3 |  |
| **Serum cotinine (ng/mL)** | 35.94 (104.97) | 36.06 (99.61) | 35.90 (106.71) | >0.9 |
| Missing | 295 | 119 | 176 |  |
| **Alcohol consumption** |  |  |  | 0.066 |
| 0-4 drinks per month | 70.56% | 74.48% | 69.20% |  |
| >4 drinks per month | 29.44% | 25.52% | 30.80% |  |
| Missing | 280 | 74 | 206 |  |
| **Waist circumference (cm)** | 101.52 (14.66) | 99.38 (13.97) | 102.30 (14.83) | <0.001 |
| Missing | 370 | 69 | 301 |  |
| **Urinary creatinine (mg/dL)** | 104.80 (67.91) | 107.46 (71.20) | 103.88 (66.72) | 0.4 |
| Missing | 139 | 44 | 95 |  |
| **Fish and seafood consumption in past 30 days** |  |  |  | 0.13 |
| 0 times | 14.38% | 15.61% | 13.93% |  |
| 1-3 times | 33.68% | 36.64% | 32.61% |  |
| 4+ times | 51.94% | 47.75% | 53.46% |  |
| Missing | 444 | 53 | 391 |  |
| **Estimated glomerular filtration rate (eGFR; ml/min/1.73 m<sup>2</sup>)** |  |  |  | <0.001 |
| eGFR < 60 | 21.33% | 11.96% | 24.52% |  |
| eGFR ≥ 60 | 78.67% | 88.04% | 75.48% |  |
| Missing | 288 | 92 | 196 |  |

<sup>1</sup>Mean (SD); %; unweighted N
<sup>2</sup>t-test adapted to complex survey samples; Chi-squared test with Rao & Scott's second-order correction Missing data for education, DSST, smoking status, cotinine, alcohol consumption, waist circumference, and creatinine imputed prior to analyses.

Our analytic sample included 147 chemicals from 14 chemical families (**Figure 1B**). Descriptive statistics including concentration minimums and maximums, means, standard deviations, medians, interquartile ranges, and the percentage of measurements above the LOD are available for all 147 chemicals (**Supplemental Table 4**). Serum cotinine was measured in the most participants (n = 4,687), followed by blood cadmium and blood lead (n = 3,955 each). Benzaldehyde had the fewest number of measurements with n = 377. Of the 147 included chemicals, 90 had detection rates of 90% or higher (**Supplemental Figure 1**).

### Associations Between Chemical Concentrations and DSST Scores

In our primary adjusted regression analysis, five of 147 exposures were associated (p < 0.01) with DSST score (**Figure 2**). The identified exposures included chemicals in the classes of metals, personal care compounds, smoking-related compounds, and volatile organic compounds. Increases in the concentrations of serum cotinine and urinary tungsten were associated with lower DSST scores (**Table 2**). A standard-deviation increase in the log_2_-transformed concentration of serum cotinine was associated with 2.71 points lower DSST (95% CI: -3.69, -1.73). Similarly, a standard-deviation increase in the log_2_-transformed concentration of urinary tungsten was associated with 1.34 points lower DSST (95% CI: -2.11, -0.56). An increase in three identified exposures (urinary benzophenone-3, blood m-/p-xylene, and urinary 2-methylhippuric acid) were associated with higher DSST scores (**Table 2**). Two exposures, serum cotinine and blood m-/p-xylene, had FDR-adjusted p-values < 0.05. Results for all screened exposures, regardless of statistical significance, are available in **Supplemental Table 5** and as a forest plot (**Supplemental Figure 2**).

**Figure 2.**
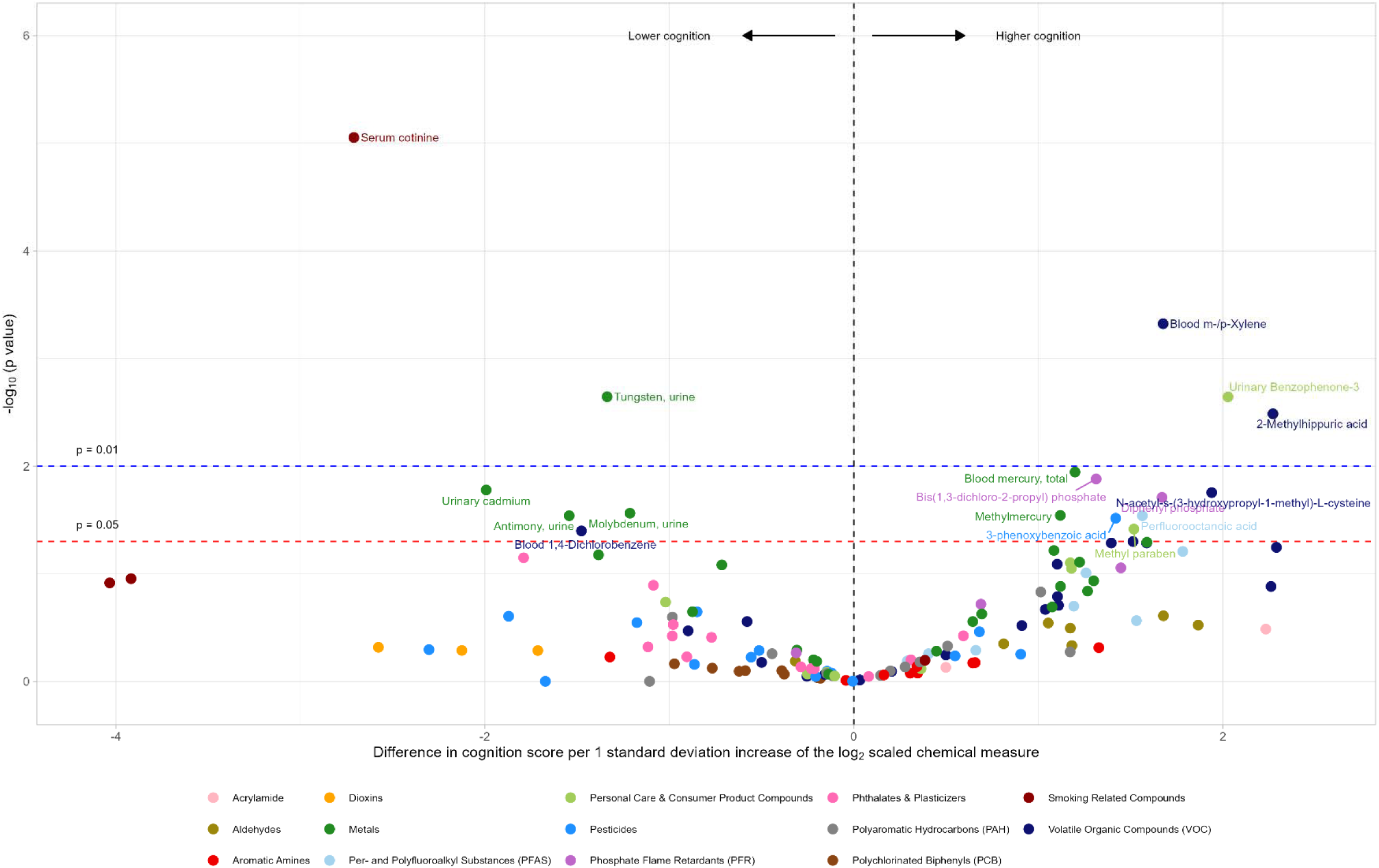
Volcano plot of associations between chemical exposures and Digit Symbol Substitution Test (DSST) scores among participants aged 60+ in the National Health and Nutrition Examination Survey (NHANES) cycles 1999-2000, 2011-2012, and 2013-2014. Beta coefficients of the chemical exposure are plotted on the x-axis and represent the difference in DSST score per 1-standard deviation increase of the log_2_-transformed exposure measure. The black vertical line indicates a beta coefficient of 0, representing no difference in DSST score. P-values are plotted on the y-axis on a -log_10_ scale, so that p-values closer to 1 are at the bottom of the plot. The red horizontal line indicates the threshold of p = 0.05 and the blue horizontal line indicates the threshold of p = 0.01.

**Table 2.** Summary of survey-weighted linear regression models evaluating the association between chemical exposure biomarkers and Digit Symbol Substitution Test (DSST) scores, NHANES 1999-2000 and 2011-2014 (overall N = 4,982), with p-values < 0.05.

| Factor | n | $\beta$ (95% CI) | p-value (unadj) | p-value (adj) |
| --- | --- | --- | --- | --- |
| **Metals** |  |  |  |  |
| Methylmercury (ug/L) | 2431 | 1.12 (0.21, 2.02) | 0.03 | 0.30 |
| Blood mercury, total (ug/L) | 2439 | 1.20 (0.39, 2.01) | 0.01 | 0.28 |
| Antimony, urine (ng/mL) | 1560 | -1.54 (-2.85, -0.23) | 0.03 | 0.30 |
| Cadmium, urine (ng/mL) | 1549 | -1.99 (-3.53, -0.46) | 0.02 | 0.29 |
| Molybdenum, urine (ng/mL) | 1549 | -1.21 (-2.22, -0.21) | 0.03 | 0.30 |
| Tungsten, urine (ng/mL) | 1573 | -1.34 (-2.11, -0.56) | 0.002 | 0.08 |
| **Per- and Polyfluoroalkyl Substances (PFAS)** |  |  |  |  |
| Perfluorooctanoic acid (ng/mL) | 1384 | 1.56 (0.23, 2.89) | 0.03 | 0.30 |
| **Personal Care & Consumer Product Compounds** |  |  |  |  |
| Benzophenone-3, urine (ng/mL) | 1134 | 2.03 (0.96, 3.09) | 0.002 | 0.08 |
| Methyl paraben (ng/mL) | 1134 | 1.52 (0.22, 2.82) | 0.04 | 0.35 |
| **Pesticides** |  |  |  |  |
| 3-phenoxybenzoic acid (ug/L) | 1047 | 1.42 (0.26, 2.58) | 0.03 | 0.30 |
| **Phosphate Flame Retardants (PFR)** |  |  |  |  |
| Bis(1,3-dichloro-2-propyl) phosphate (ug/L) | 957 | 1.31 (0.40, 2.23) | 0.01 | 0.28 |
| Diphenyl phosphate (ug/L) | 967 | 1.67 (0.43, 2.92) | 0.02 | 0.29 |
| **Smoking Related Compounds** |  |  |  |  |
| Serum cotinine (ng/mL) | 4687 | -2.71 (-3.69, -1.73) | $8.9 \times 10^{-6}$ | 0.001 |
| **Volatile Organic Compounds (VOC)** |  |  |  |  |
| Blood 1,4-Dichlorobenzene (ng/mL) | 1475 | -1.48 (-2.77, -0.19) | 0.04 | 0.35 |
| m-/p-Xylene, blood (ng/ml) | 1442 | 1.68 (0.92, 2.43) | $4.8 \times 10^{-4}$ | 0.03 |
| 2-Methylhippuric acid, urine (ng/mL) | 1022 | 2.27 (1.00, 3.55) | 0.003 | 0.10 |
| N-acetyl-s-(3-hydroxypropyl-1-methyl)-L-cysteine (ng/mL) | 1068 | 1.94 (0.51, 3.37) | 0.02 | 0.29 |

Models adjusted for: age, sex, race/ethnicity, education, smoking status, blood cotinine, seafood consumption, NHANES cycle, and urinary creatinine (urinary measures only)
Chemical exposure, blood cotinine, and urinary creatinine were log<sub>2</sub> transformed and z-score standardized
P-values adjusted by controlling for the False Discovery Rate (FDR)

### Sensitivity Analyses

When restricting the analytic group to participants with normal kidney function (N = 3,593), three chemical biomarkers were associated (p < 0.01) with DSST score (**Supplemental Figure 3** and **Supplemental Table 6**). All identified associations were consistent with the results of the main analysis.

When adjusting further for waist circumference and alcohol consumption in the full analytic sample, the previously identified associations between blood mercury and 2-methylhippuric acid and DSST were no longer significant (**Supplemental Figure 4** and **Supplemental Table 7**). Notably, an increase in urinary cadmium concentration was associated with a decrease in DSST score by 2.72 points (95% CI: -4.27, -1.18).

After MICE, the 25^th^ percentile of DSST score used to define the cutoff for MCI was 29. A total of five biomarkers were associated (p < 0.01) with MCI in modified Poisson regression models (**Figure 3** and **Table 3**). Consistent with the continuous results, serum cotinine was associated with higher prevalence of MCI (RR 1.18, 95% CI: 1.07, 1.31). Mono-benzyl phthalate was also associated with higher prevalence of MCI (RR 1.23, 95% CI: 1.07, 1.41). Additionally, 3 biomarkers were associated with lower prevalence of mild cognitive impairment: 2-methylhippuric acid (RR 0.71, 95% CI: 0.61, 0.83), combined 3- and 4-methylhippuric acid (RR 0.74, 95% CI: 0.62, 0.89), and perfluorooctanoic acid (PFOA) (RR 0.81, 95% CI: 0.70, 0.93). Poisson regression results for all exposures are available in **Supplemental Table 8**.

**Figure 3.**
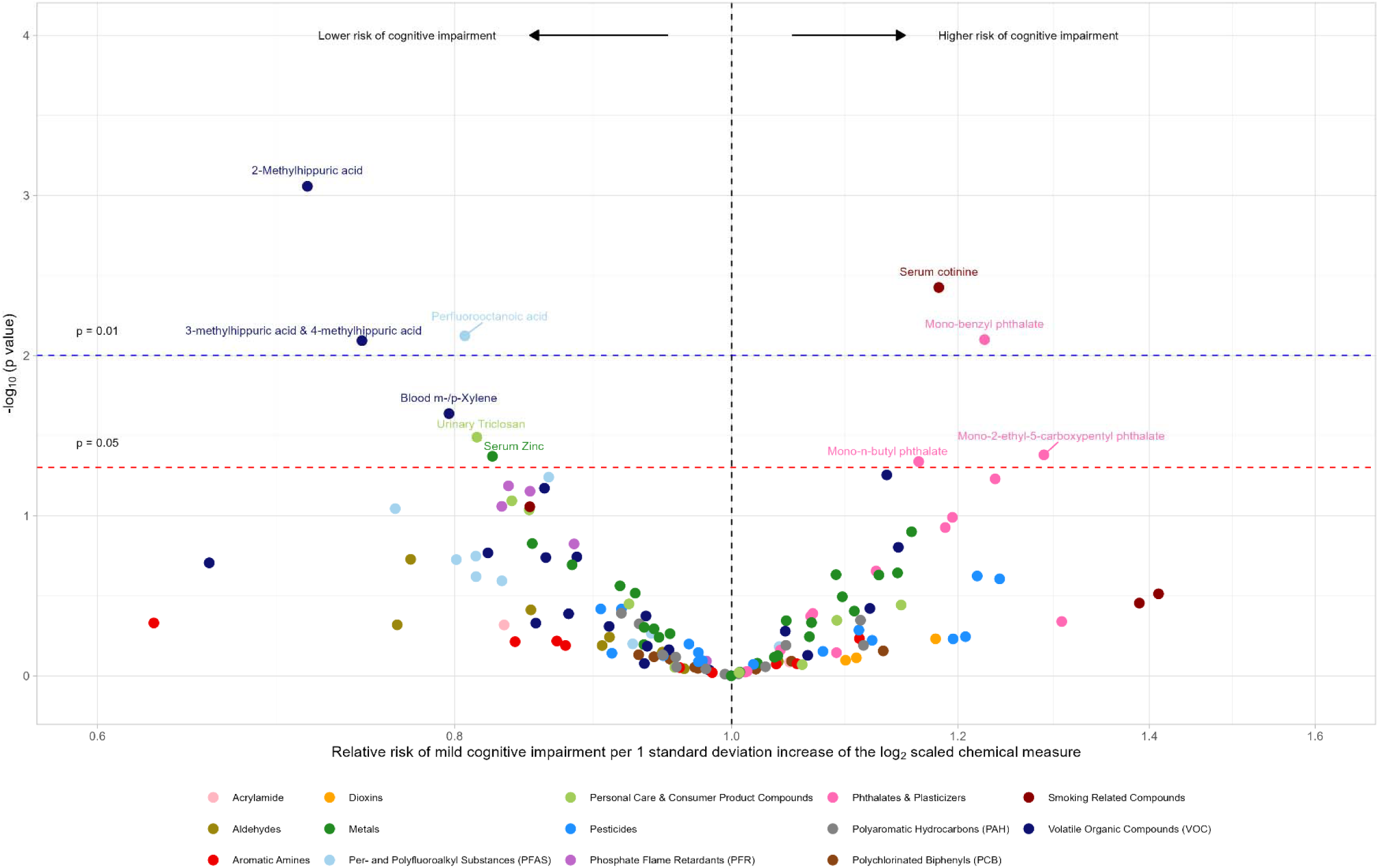
Volcano plot of associations between chemical exposures and mild cognitive impairment (MCI) among participants aged 60+ in the National Health and Nutrition Examination Survey (NHANES) cycles 1999-2000, 2011-2012, and 2013-2014. Relative risks (prevalence ratios) derived from modified Poisson regression models are plotted on the x-axis and represent a 1-standard deviation increase of the log_2_-transformed exposure measure. The black vertical line indicates a relative risk of 1, representing no association with MCI. P-values are plotted on the y-axis on a -log_10_ scale, so that p-values closer to 1 are at the bottom of the plot. The red horizontal line indicates the threshold of p = 0.05 and the blue horizontal line indicates the threshold of p = 0.01.

**Table 3.** Summary of survey-weighted modified Poisson regression models evaluating the association between chemical exposure biomarkers and binary mild cognitive impairment status, NHANES 1999-2000 and 2011-2014 (overall N = 4,982), with p-values < 0.05.

| Factor | n | Prevalence ratio (95% CI) | p-value (unadj) | p-value (adj) |
| --- | --- | --- | --- | --- |
| **Metals** |  |  |  |  |
| Serum zinc (ug/dL) | 1065 | 0.82 (0.70, 0.98) | 0.04 | 0.64 |
| **Per- and Polyfluoroalkyl Substances** |  |  |  |  |
| Perfluorooctanoic acid, blood (ng/mL) | 1384 | 0.81 (0.70, 0.93) | 0.008 | 0.24 |
| **Personal Care & Consumer Product Compounds** |  |  |  |  |
| Urinary triclosan (ng/mL) | 1134 | 0.81 (0.69, 0.96) | 0.03 | 0.64 |
| **Phthalates & Plasticizers** |  |  |  |  |
| Mono-2-ethyl-5-carboxypentyl phthalate (ng/mL) | 1134 | 1.29 (1.04, 1.59) | 0.04 | 0.64 |
| Mono-benzyl phthalate, urine (ng/mL) | 1647 | 1.23 (1.07, 1.41) | 0.008 | 0.24 |
| Mono-n-butyl phthalate (ng/mL) | 1647 | 1.16 (1.01, 1.34) | 0.046 | 0.64 |
| **Smoking Related Compounds** |  |  |  |  |
| Cotinine, blood (ng/mL) | 4687 | 1.18 (1.07, 1.31) | 0.004 | 0.24 |
| **Volatile Organic Compounds (VOC)** |  |  |  |  |
| Blood m-/p-Xylene (ng/mL) | 1442 | 0.80 (0.67, 0.95) | 0.02 | 0.56 |
| 2-Methylhippuric acid, urine (ng/mL) | 1022 | 0.71 (0.61, 0.83) | 8.8x10<sup>-4</sup> | 0.13 |
| 3-methylhippuric acid & 4-methylhippuric acid, urine (ng/mL) | 1070 | 0.74 (0.62, 0.89) | 0.008 | 0.24 |

Models adjusted for: age, sex, race/ethnicity, education, smoking status, blood cotinine, seafood consumption, NHANES cycle, urinary creatinine (urinary measures only)
Chemical exposure, blood cotinine, and urinary creatinine were log<sub>2</sub> transformed and z-score standardized
P-values adjusted by controlling for the False Discovery Rate (FDR)

When restricted to participants with normal kidney function, two biomarkers continued to be associated (p < 0.01) with MCI (**Supplemental Figure 5** and **Supplemental Table 9**). The results for cotinine (RR 1.25, 95% CI: 1.07, 1.47) and PFOA (RR 0.70, 95% CI: 0.56, 0.88) were consistent with the full sample results.

## Discussion

In a large, diverse, and nationally representative sample of older adults (ages 60+) in the United States, we investigated the cross-sectional relationship between chemical exposure biomarkers and cognition. We used an ExWAS framework to separately and systematically examine 147 chemicals. We observed that elevated levels of cotinine, cadmium, and tungsten were associated with lower cognitive measures and mono-benzyl phthalate was associated with greater risk of cognitive impairment. On the other hand, we observed that higher biomarker concentrations of benzophenone-3, m-/p-xylene, and 2-methylhippuric acid were associated with higher cognitive scores. Similarly, PFOA and combined 3-methylhippuric acid/4-methylhippuric acid were associated with lower risk of cognitive impairment. These hypothesis-generating findings broaden the landscape of chemicals for testing with neurodegenerative outcomes, and these chemicals may be modifiable prevention factors.

Our findings are consistent with the literature in several respects. We observed cotinine levels were associated with lower cognition. Cotinine itself is a nicotine metabolite that weakly activates nicotinic acetylcholine receptors.^36^ More broadly, cotinine is a biomarker with high sensitivity and specificity for recent (3-4 days) cigarette smoke exposure.^37^ Cigarette smoke exposure is well recognized as a risk factor for cognitive impairment and dementia, and cessation even in later life is preventative.^5^ Smoking in later life is estimated to account for 5% of dementia cases.^5^ In epidemiologic studies, it can be challenging to study the relationship between cigarette smoke exposure and dementia due to mortality from competing risks of other health effects such as lung cancer and cardiovascular disease that may occur at younger ages.^38^ Cigarette smoke is a complex exposure mixture of over 9,500 compounds, including metals, gases, and particulate matter.^39^ Particulate matter is well-replicated as a dementia risk factor.^40,41^ Inhalation of smoking-related compounds results in absorption and toxicity in the lungs. The lungs are also highly vascularized, and these compounds can have systemic vascular impacts, which can impact cognition.^42^ These compounds can also circulate in the bloodstream and cross the blood-brain-barrier for direct neurotoxic effects.^43^ Our findings related to cotinine and cognition align with the current toxicologic and epidemiologic literature.

We also observed that urinary cadmium was associated with lower cognition in a sensitivity analysis. Cadmium is a metal with no normal physiologic function in the body.^44^ The primary cadmium exposure routes are through cigarette smoke (tobacco plants bioaccumulate cadmium from the soil) and diet (rice, leafy green vegetables, grains).^45,46^ Our findings were robust to adjustment for potential confounding due to cigarette smoke exposure (through self-reported smoking status and cotinine levels) and fish consumption. Cadmium exposure has neurotoxic effects in animal models and in cell culture.^47–49^ In human studies, cadmium has previously been associated with impaired cognition or dementia. Specifically, in NHANES (1999-2006, n=2,023) data linked to the National Death Index, an interquartile range increase (0.78 ng/mL) in urinary cadmium was associated with 1.58 times higher hazard of Alzheimer’s disease mortality over 7.5 years of follow up.^6^ Similarly, among 4,064 NHANES participants (1999-2004) the highest quartile of blood cadmium (>0.6ug/L) was associated with 3.76 times higher hazard of Alzheimer’s disease mortality relative to the lowest quartile.^50^ Notably, these studies were prospective and showed cadmium exposure preceded Alzheimer’s mortality by several years. Our findings, though cross-sectional, are consistent with this direction of association.

One of the new findings in our study is that elevated tungsten levels were associated with lower cognition. Tungsten is a dense metal that is widely used in industrial applications due to its thermal, electrical, and anti-corrosive properties.^51^ It replaced lead in bullets and is detected in environmental samples near military and mining sites.^52^ A primary occupational route of exposure is inhalation.^52^ Lung cell models have demonstrated tungsten toxicity with implications for tumorigenesis.^53,54^ Pulmonary, cardiometabolic, bone, and immune toxicity have also been noted with tungsten exposure.^55^ To date, little research has been conducted on tungsten neurotoxicity. A previous NHANES analysis (2011-2014, n=888) of multiple metals and cognition observed log-transformed urinary tungsten levels were associated with lower cognition on the Consortium to Establish a Registry for Alzheimer’s Disease (CERAD) immediate (-0.38 points, 95% CI: -0.75, -0.01) and delayed (-0.19 points, 95% CI: -0.38, -0.004) recall tests.^56^ Because the CERAD test was only available in two NHANES waves, we elected to focus on the DSST measures. Nonetheless, the direction of tungsten association in this previous study is consistent with our findings.

The present ExWAS approach also discovered that higher concentrations of certain biomarkers were linked to better cognition scores or lower prevalences of cognitive impairment. Some of these findings were unexpected, and we cannot rule out the possibility that they occurred by chance. The findings for mercury and PFOA were not surprising as similar associations have been reported.^57–61^ One potential explanation is the presence of negative confounding. Fish consumption, a major source of methylmercury and PFAS,^62,63^ is also known to protect against memory loss and neurodegeneration.^64^ Kidney function is another critical factor as poor kidney function can influence biomarker concentrations.^34^ Poor kidney function has also been associated with worse cognitive function.^35^ To address the potential impact of these factors, we adjusted for fish consumption and conducted a sensitivity analysis restricted to normal kidney function,^62,65^ but the associations remained unchanged. We were also surprised that well documented neurotoxicants, such as lead^7^, were not associated with cognition in our study. Perhaps the exposure timing in our study did not align with the window of susceptibility of lead exposure’s impacts on cognition, or the exposure distribution was too narrow to be adequately powered to detect an association. These findings require further investigation through a targeted approach using a carefully designed study.

Limitations of this study point to potential future directions. Most notably, the design was cross-sectional (exposure and outcome measured at the same time point). This limits our ability to understand the temporal sequence between exposure and outcome and findings may be subject to reverse causation. For example, while elevated chemical exposure levels may be risk factors for cognitive impairment, it is also possible that prevalent cognitive impairment alters participant behaviors, which in turn alters chemical exposure levels. Findings from this cross-sectional discovery analysis will allow for follow up studies to examine a more targeted list of chemicals in a longitudinal or prospective design. In the NHANES design, chemical measures are not measured on all participants. While our analyses captured all available participants with measures, for some chemicals, the analytic sample size was quite limited. We may have been underpowered to detect associations for these chemicals. It will be important to test for replication and meta-analyze with additional studies. In the ExWAS framework, chemicals are analyzed with the same regression model. In our case, we elected to use a log-linear model, which is a common dose response curve in cognitive studies.

However, for a given chemical the true dose-response curve may differ (linear, exponential, etc.). Follow up studies of individual chemicals may be able to explore additional model forms, which may better fit the specific data. We also evaluated chemicals individually, though their exposure levels may be correlated, and they may occur in complex mixtures.^66^ Additional analyses may consider mixtures analyses.^67^ Mixtures analyses were not implemented in the current paper because the research question related to chemical discovery, which is more appropriately addressed in the

ExWAS framework. Logistical considerations in this study also impeded the implementation of potential mixtures analyses, including minimal overlap of participants across exposure measures. Another potential limitation relates to the specificity of the DSST, which requires participant vision, physical mobility, and coordination for completion. Lower scores on this DSST may not purely reflect cognitive function but also other impairments. Lastly, though we accounted for several common confounders of exposures and cognition (smoking status, cotinine levels, fish consumption, kidney function, alcohol consumption, waist circumference), it is still possible that we have unmeasured confounding between exposures and cognition. We think this is particularly likely for chemicals whose exposure levels are associated with increased cognition, and for chemicals which these relationships do not have a biologic plausibility. Future studies of individual chemicals may be able to explore additional confounders or mechanisms.

This study had key strengths. First, the study population was diverse and nationally representative to the United States. This is important for generalizability of findings. Analytically, we accounted for sample weights and clustering strategies. Second, the analysis included assessment of many chemicals from several chemical classes. Examining a wide array of chemicals is important for hypothesis generation. These chemicals were all measured in either blood or urine using rigorous standards. Third, the cognition measure used in this study is broadly applicable and has been implemented in other settings, enabling comparability across studies. For greatest statistical power we examined continuous cognition and for clinical translation we examined cognition dichotomized by impairment status. Fourth, we used current ExWAS analytic standards to test and visualize associations and account for multiple comparisons.^10^ We also used multiple imputation approaches to account for missing data in covariate and outcome measures, so that we retained the largest possible sample size. Fifth, we performed several sensitivity analyses which improved the robustness of our findings, including restricting on kidney function status, as some chemicals are metabolized by renal tissues, influencing measured biomarker concentration levels. Together, this study advances the field of environmental epidemiology of cognition among older adults.

In summary, this study examined the independent relationships between 147 chemicals with cross-sectional cognition in a large and nationally representative sample of 4,982 adults over age 60 in the United States. We confirmed previous observations that elevated levels of cotinine and cadmium are associated with lower cognition. Using this discovery approach, we also identified novel associations between tungsten and mono-benzyl phthalate and cognition. These findings can inspire new laboratory toxicologic investigations as well as longitudinal epidemiologic studies to examine the windows of chemical susceptibility. This paper adds to a building scholarship on environmental risk factors for impaired cognition among older adults. Identifying environmental contributors to Alzheimer’s disease and related dementias is part of the 2023 updated National Plan to Address Alzheimer’s Disease.^68^ Together, these studies offer opportunities for public health prevention through policy and practice to reduce chemical exposures and disparities.

## Data Availability

All data referred to in the manuscript are publicly available (raw data: https://www.cdc.gov/nchs/nhanes/index.htm; processed data: https://www.kaggle.com/datasets/nguyenvy/nhanes-19882018). All code to analyze the data are publicly available (https://github.com/bakulskilab/Cognition_ExWAS).

https://www.cdc.gov/nchs/nhanes/index.htm

https://www.kaggle.com/datasets/nguyenvy/nhanes-19882018

## Acronyms

DSST: Digit Symbol Substitution Test
ExWAS: Exposome-Wide Association Study
FDR: False Discovery Rate
GED: General Education Development
IRB: Institutional Review Board
LOD: Limit of Detection
MCI: Mild Cognitive Impairment
MICE: Multiple Imputation by Chained Equations
NCHS: National Center for Health Statistics
NHANES: National Health and Nutrition Examination Survey
US: United States

## Acknowledgements

This research was supported by funds from the National Institute on Aging (P30 AG072931, R01 AG070897, U01 AG009740-33S5, R01 AG072396, R01 AG067592), the National Institute of Environmental Health Sciences (P30 ES017885), and the National Institute of General Medical Sciences (T32 GM007863-41).

## Notes

### Competing Interest Statement

The authors have declared no competing interest.

### Author Declarations

All source data were openly available before the start of this study and can be accessed through the National Center for Health Statistics (https://www.cdc.gov/nchs/nhanes/index.htm).

