## Supplement for "Exposome-wide association study of cognition among older adults in the National Health and Nutrition Examination Survey"

**Supplemental Material**

This document contains:

5 Supplemental figures

9 Supplemental tables

| **Supplemental Table 1.** Summary of analytic variables with missing data imputed using Multiple Imputation by Chained Equations (MICE). | | |
| --- | --- | --- |
| **Variable** | **N missing** | **Imputation method** |
| Education level | 15 | Proportional odds modeling |
| Urinary creatinine (mg/dL) | 139 | Predictive mean modeling |
| Waist circumference (cm) | 370 | Predictive mean modeling |
| Serum cotinine (ng/mL) | 295 | Predictive mean modeling |
| DSST score | 721 | Predictive mean modeling |
| Smoking status | 7 | Polytomous logistic regression |
| Seafood consumption (number of times in past month) | 444 | Predictive mean modeling |
| Alcohol consumption (drinks per month) | 280 | Predictive mean modeling |

**Supplemental Table 2.** Weighted descriptive statistics of excluded vs. included participants aged 60 years and older, National Health and Nutrition Examination Survey (NHANES) 1999-2000, 2011-2012, and 2013-2014 (N = 5,466).

| **Variable** | **Overall**  **(N = 5,466)**^1^ | **Excluded**  **(N = 484)**^1^ | **Included**  **(N = 4,982)**^1^ | **p-value**^2^ |
| --- | --- | --- | --- | --- |
| **Digit Symbol Substitution Test (DSST)** | 50.30 (17.41) | 36.60 (14.40) | 50.42 (17.38) | <0.001 |
| Missing | 1,035 | 314 | 721 |  |
| **Age (years)** | 69.82 (6.99) | 72.10 (7.79) | 69.79 (6.98) | 0.025 |
| **Sex** |  |  |  | >0.9 |
| Male | 44.98% | 44.48% | 44.98% |  |
| Female | 55.02% | 55.52% | 55.02% |  |
| **Race/ethnicity** |  |  |  | <0.001 |
| Mexican American | 3.59% | 2.00% | 3.61% |  |
| Other Hispanic | 4.60% | 5.79% | 4.59% |  |
| Non-Hispanic White | 78.05% | 65.43% | 78.23% |  |
| Non-Hispanic Black | 8.73% | 23.26% | 8.52% |  |
| Other Race | 5.03% | 3.52% | 5.05% |  |
| **Education** |  |  |  | 0.3 |
| Did not complete high school | 23.08% | 28.94% | 23.00% |  |
| Completed high school or above | 76.92% | 71.06% | 77.00% |  |
| Missing | 24 | 9 | 15 |  |
| **Smoking status** |  |  |  | 0.5 |
| Never smoker | 49.16% | 55.92% | 49.06% |  |
| Former smoker | 39.10% | 34.94% | 39.16% |  |
| Current smoker | 11.74% | 9.15% | 11.78% |  |
| Missing | 14 | 7 | 7 |  |
| **Serum cotinine (ng/mL)** | 35.94 (104.97) | NA | 35.94 (104.97) |  |
| Missing | 779 | 484 | 295 |  |
| **Alcohol consumption** |  |  |  | 0.066 |
| 0-4 drinks per month | 70.70% | 85.00% | 70.56% |  |
| >4 drinks per month | 29.30% | 15.00% | 29.44% |  |
| Missing | 701 | 421 | 280 |  |
| **Waist circumference (cm)** | 101.54 (14.67) | 105.15 (16.24) | 101.52 (14.66) | 0.2 |
| Missing | 809 | 439 | 370 |  |
| **Urinary creatinine (mg/dL)** | 104.78 (67.95) | 102.38 (74.37) | 104.80 (67.91) | 0.9 |
| Missing | 578 | 439 | 139 |  |
| **Fish and seafood consumption in past 30 days** |  |  |  | 0.3 |
| 0 times | 14.47% | 24.18% | 14.38% |  |
| 1-3 times | 33.72% | 37.85% | 33.68% |  |
| 4+ times | 51.81% | 37.97% | 51.94% |  |
| Missing | 866 | 422 | 444 |  |
| **Estimated glomerular filtration rate (eGFR; ml/min/1.73 m2)** |  |  |  |  |
| eGFR < 60 | 21.33% | NA | 21.33% |  |
| eGFR >= 60 | 78.67% | NA | 78.67% |  |
| Missing | 772 | 484 | 288 |  |
| **NHANES cycle** |  |  |  | <0.001 |
| 1 | 25.92% | 46.00% | 25.64% |  |
| 7 | 35.91% | 43.62% | 35.80% |  |
| 8 | 38.17% | 10.38% | 38.56% |  |
| ^1^Mean (SD); %; unweighted N missing | | | | |
| ^2^t-test adapted to complex survey samples; chi-squared test with Rao & Scott's second-order correction | | | | |

| **Supplemental Table 3.** Weighted descriptive statistics of participants, stratified by cognitive impairment status determined by Digit Symbol Substitution Test (DSST) score, in the National Health and Nutrition Examination Survey (NHANES) 1999-2000, 2011-2012, and 2013-2014 (N = 4,982). | | | | |
| --- | --- | --- | --- | --- |
| **Variable** | **Mild cognitive impairment  (N = 889)^1^** | **No cognitive impairment  (N = 3,372)^1^** | **Missing DSST score  (N = 721)^1^** | **p-value^2^** |
| **Digit Symbol Substitution Test (DSST)** | 19.72 (7.12) | 54.23 (14.20) | NA | <0.001 |
| Missing | 0 | 0 | 721 |  |
| **Age (years)** | 73.33 (7.01) | 68.95 (6.66) | 73.11 (7.38) | <0.001 |
| **Sex** |  |  |  | 0.6 |
| Male | 44.59% | 45.34% | 42.48% |  |
| Female | 55.41% | 54.66% | 57.52% |  |
| **Race/ethnicity** |  |  |  | <0.001 |
| Mexican American | 9.42% | 2.44% | 7.30% |  |
| Other Hispanic | 14.88% | 2.93% | 7.70% |  |
| Non-Hispanic White | 49.39% | 84.08% | 59.60% |  |
| Non-Hispanic Black | 21.57% | 6.05% | 15.48% |  |
| Other Race | 4.74% | 4.49% | 9.92% |  |
| **Education** |  |  |  | <0.001 |
| Did not complete high school | 60.07% | 15.22% | 49.42% |  |
| Completed high school or above | 39.93% | 84.78% | 50.58% |  |
| Missing | 3 | 3 | 9 |  |
| **Smoking status** |  |  |  | 0.047 |
| Never smoker | 52.23% | 48.18% | 53.12% |  |
| Former smoker | 35.04% | 40.44% | 32.89% |  |
| Current smoker | 12.73% | 11.39% | 13.99% |  |
| Missing | 0 | 4 | 3 |  |
| **Serum cotinine (ng/mL)** | 45.83 (119.74) | 32.64 (98.09) | 54.32 (138.72) | <0.001 |
| Missing | 68 | 153 | 74 |  |
| **Alcohol consumption** |  |  |  | <0.001 |
| 0-4 drinks per month | 86.90% | 67.10% | 86.97% |  |
| >4 drinks per month | 13.10% | 32.90% | 13.03% |  |
| Missing | 48 | 61 | 171 |  |
| **Waist circumference (cm)** | 101.26 (14.10) | 101.60 (14.60) | 100.95 (15.97) | 0.4 |
| Missing | 79 | 125 | 166 |  |
| **Urinary creatinine (mg/dL)** | 112.65 (72.64) | 103.96 (67.56) | 103.70 (65.14) | 0.079 |
| Missing | 28 | 36 | 75 |  |
| **Fish and seafood consumption in past 30 days** |  |  |  | <0.001 |
| 0 times | 25.12% | 12.79% | 17.90% |  |
| 1-3 times | 42.46% | 31.93% | 41.21% |  |
| 4+ times | 32.42% | 55.27% | 40.89% |  |
| Missing | 85 | 192 | 167 |  |
| **Estimated glomerular filtration rate (eGFR; ml/min/1.73 m^2^)** |  |  |  | <0.001 |
| eGFR < 60 | 35.46% | 18.49% | 31.31% |  |
| eGFR >= 60 | 64.54% | 81.51% | 68.69% |  |
| Missing | 62 | 153 | 73 |  |
| **NHANES cycle** |  |  |  | <0.001 |
| 1999-2000 | 35.76% | 23.32% | 34.31% |  |
| 2011-2012 | 29.72% | 36.43% | 36.86% |  |
| 2013-2014 | 34.51% | 40.25% | 28.83% |  |
| ^1^Mean (SD); %; unweighted N | | | | |
| ^2^Kruskal-Wallis rank-sum test for complex survey samples; chi-squared test with Rao & Scott's second-order correction  Missing data for education, DSST, smoking status, cotinine, alcohol consumption, waist circumference, and creatinine imputed prior to analyses. | | | | |

**Supplemental Table 4.** Distributions of included environmental chemical biomarker levels.

Available as a separate excel file: “Supplemental Table 4.xlsx”

**Supplemental Figure 1.** Scatterplot of the sample size per chemical versus the percentage of measurements above the lower limit of detection (LOD). The points are colored by family group, and outlier points are labeled with the chemical names.

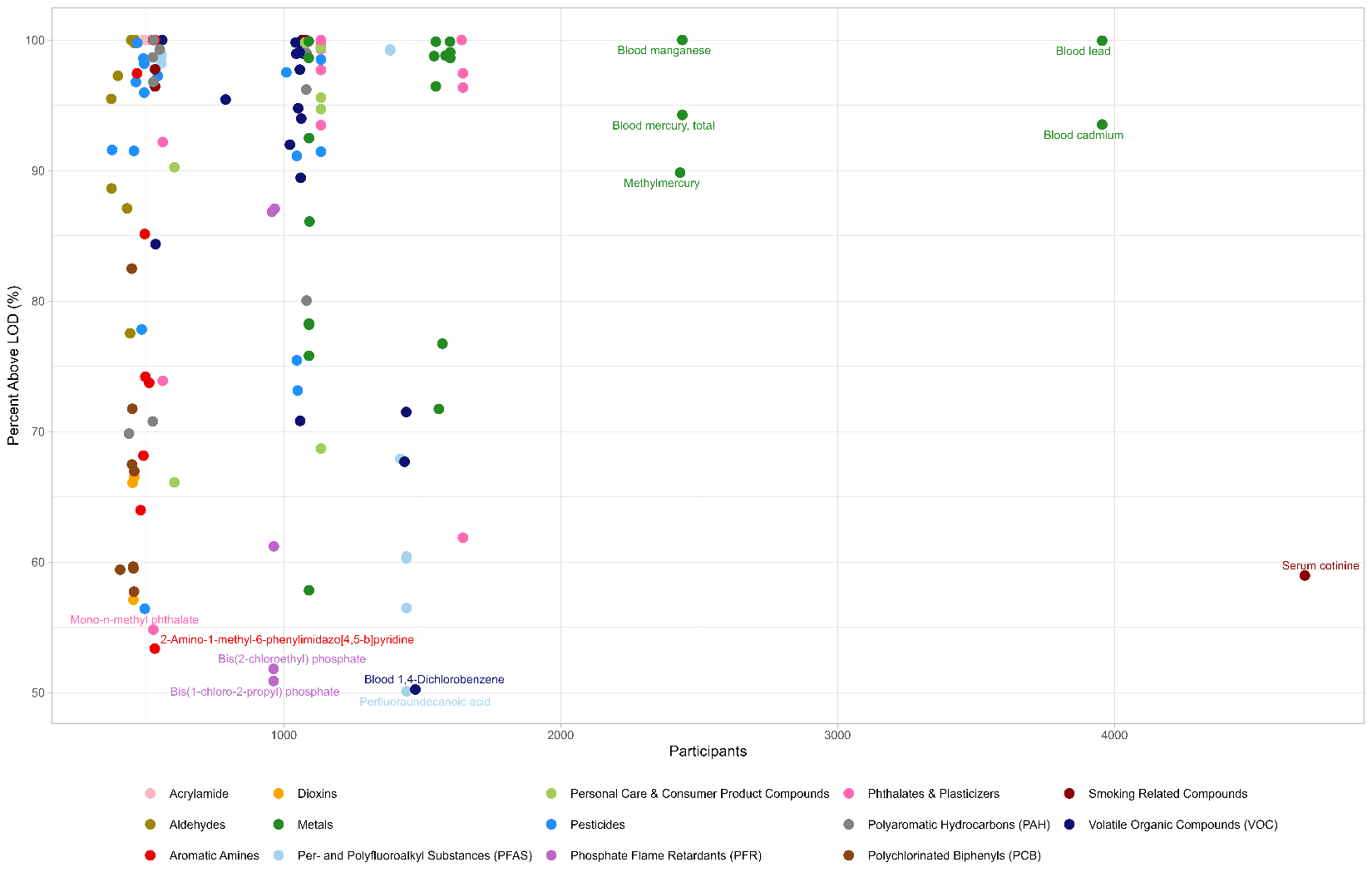

**Supplemental Figure 2.** Forest plot of beta coefficients and 95% confidence intervals from survey-weighted generalized linear models for each chemical exposure. Points are colored by chemical family. Points to the left of zero represent chemicals in which higher concentrations were associated with lower Digit Symbol Substitution Test (DSST) scores, while points to the right of zero represent chemicals in which higher concentrations were associated with higher DSST scores.

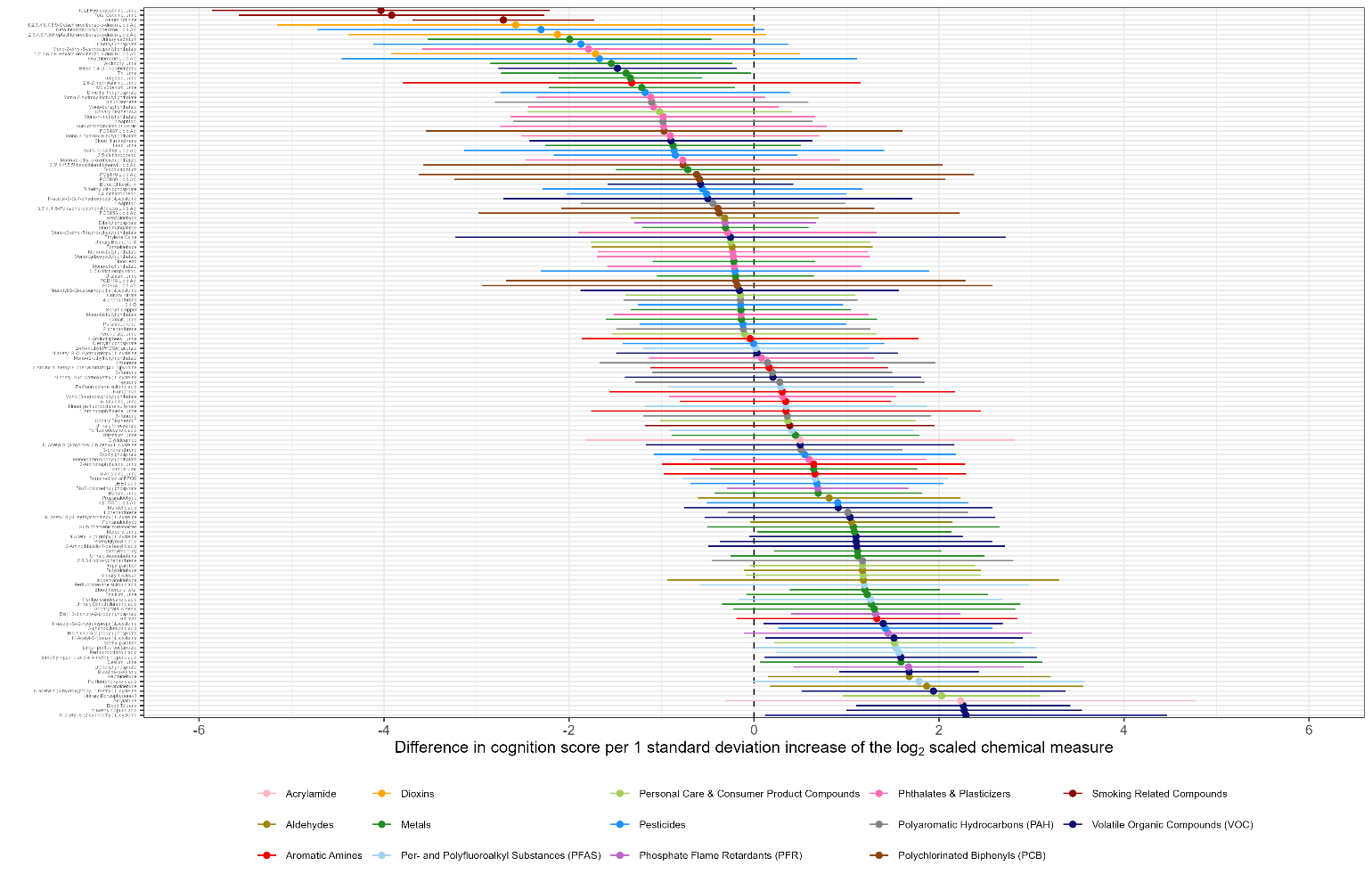

**Supplemental Table 5.** Full results for the primary analysis. Summary of survey-weighted linear regression models evaluating the association between chemical exposure biomarkers and Digit Symbol Substitution Test (DSST) scores, NHANES 1999-2000 and 2011-2014 (overall N = 4,982) for all chemicals.

Models were adjusted for age, sex, race/ethnicity, education, smoking status, blood cotinine, seafood consumption, NHANES cycle, and urinary creatinine (urinary measures only). Chemical exposure, blood cotinine, and urinary creatinine were log2 transformed and z-score standardized. P-values adjusted by controlling for the False Discovery Rate (FDR).

Available as a separate excel file: “Supplemental Table 5.xlsx”

**Supplemental Figure 3.** Volcano plot of associations between chemical exposures and Digit Symbol Substitution Test (DSST) scores among participants aged 60+ with estimated glomerular filtration rate >= 60 ml/min/1.73 m^2^ in the National Health and Nutrition Examination Survey (NHANES) cycles 1999-2000, 2011-2012, and 2013-2014. Beta coefficients of the chemical exposure are plotted on the x-axis and represent the difference in DSST score per 1-standard deviation increase of the log_2_-transformed exposure measure. The black vertical line indicates a beta coefficient of 0, representing no difference in DSST score. P-values are plotted on the y-axis on a -log_10_ scale, so that p-values closer to 1 are at the bottom of the plot. The red horizontal line indicates the threshold of p = 0.05 and the blue horizontal line indicates the threshold of p = 0.01.

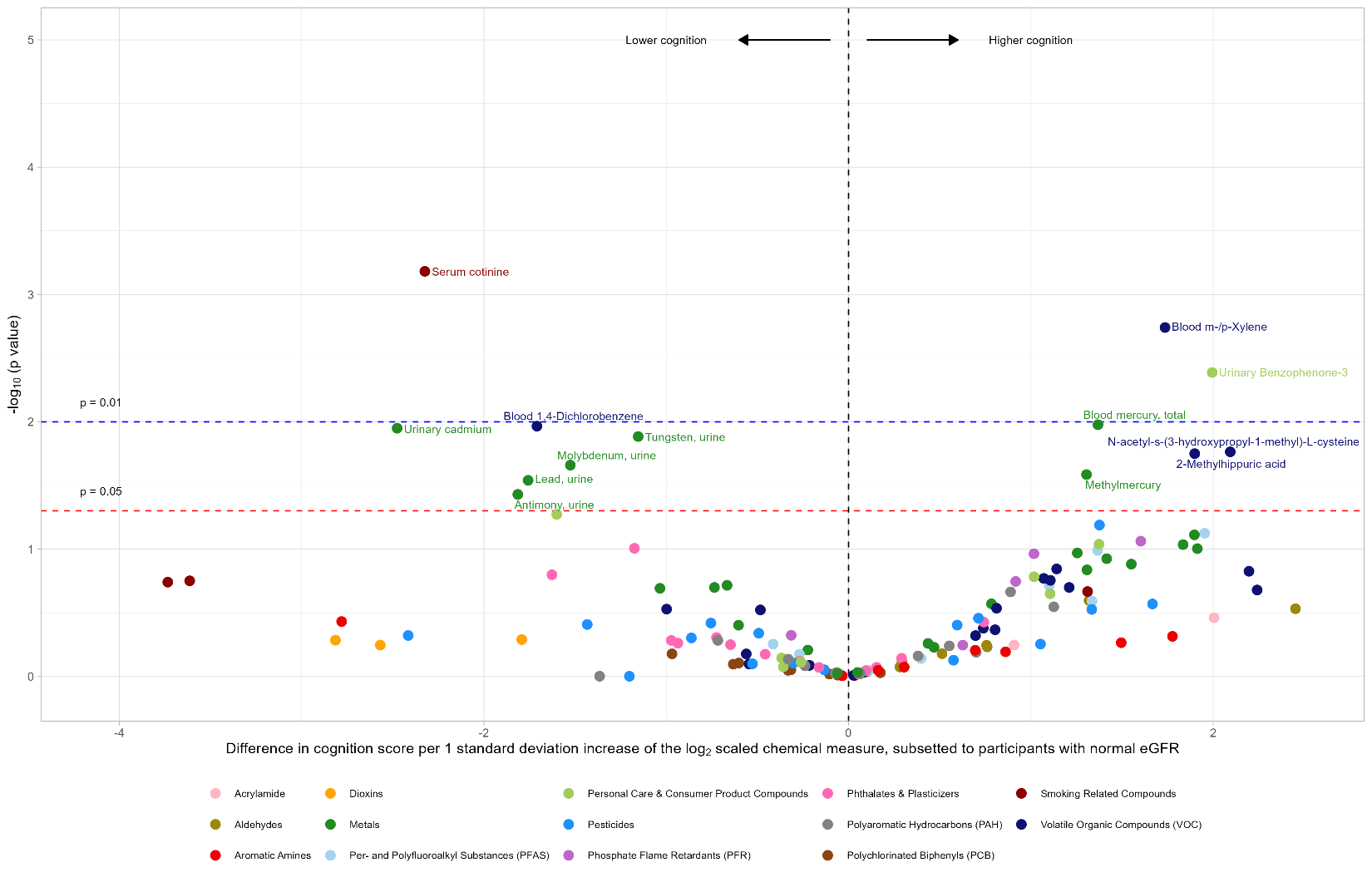

**Supplemental Table 6**. Primary model in the subset of participants with normal estimated glomerular filtration rate (eGFR, ≥60 ml/min/1.73 m^2^) as a marker of kidney function. Summary of survey-weighted linear regression models evaluating the association between chemical exposure biomarkers and Digit Symbol Substitution Test (DSST) scores, NHANES 1999-2000 and 2011-2014 (normal eGFR N = 3,593) for all chemicals.

Models were adjusted for age, sex, race/ethnicity, education, smoking status, blood cotinine, seafood consumption, NHANES cycle, and urinary creatinine (urinary measures only). Chemical exposure, blood cotinine, and urinary creatinine were log2 transformed and z-score standardized. P-values adjusted by controlling for the False Discovery Rate (FDR).

Available as a separate excel file: “Supplemental Table 6.xlsx”

**Supplemental Figure 4.** Volcano plot of associations between chemical exposures and Digit Symbol Substitution Test (DSST) scores among participants aged 60+ in the National Health and Nutrition Examination Survey (NHANES) cycles 1999-2000, 2011-2012, and 2013-2014, additionally adjusted for waist circumference and alcohol consumption. Beta coefficients of the chemical exposure are plotted on the x-axis and represent the difference in DSST score per 1-standard deviation increase of the log_2_-transformed exposure measure. The black vertical line indicates a beta coefficient of 0, representing no difference in DSST score. P-values are plotted on the y-axis on a -log_10_ scale, so that p-values closer to 1 are at the bottom of the plot. The red horizontal line indicates the threshold of p = 0.05 and the blue horizontal line indicates the threshold of p = 0.01.

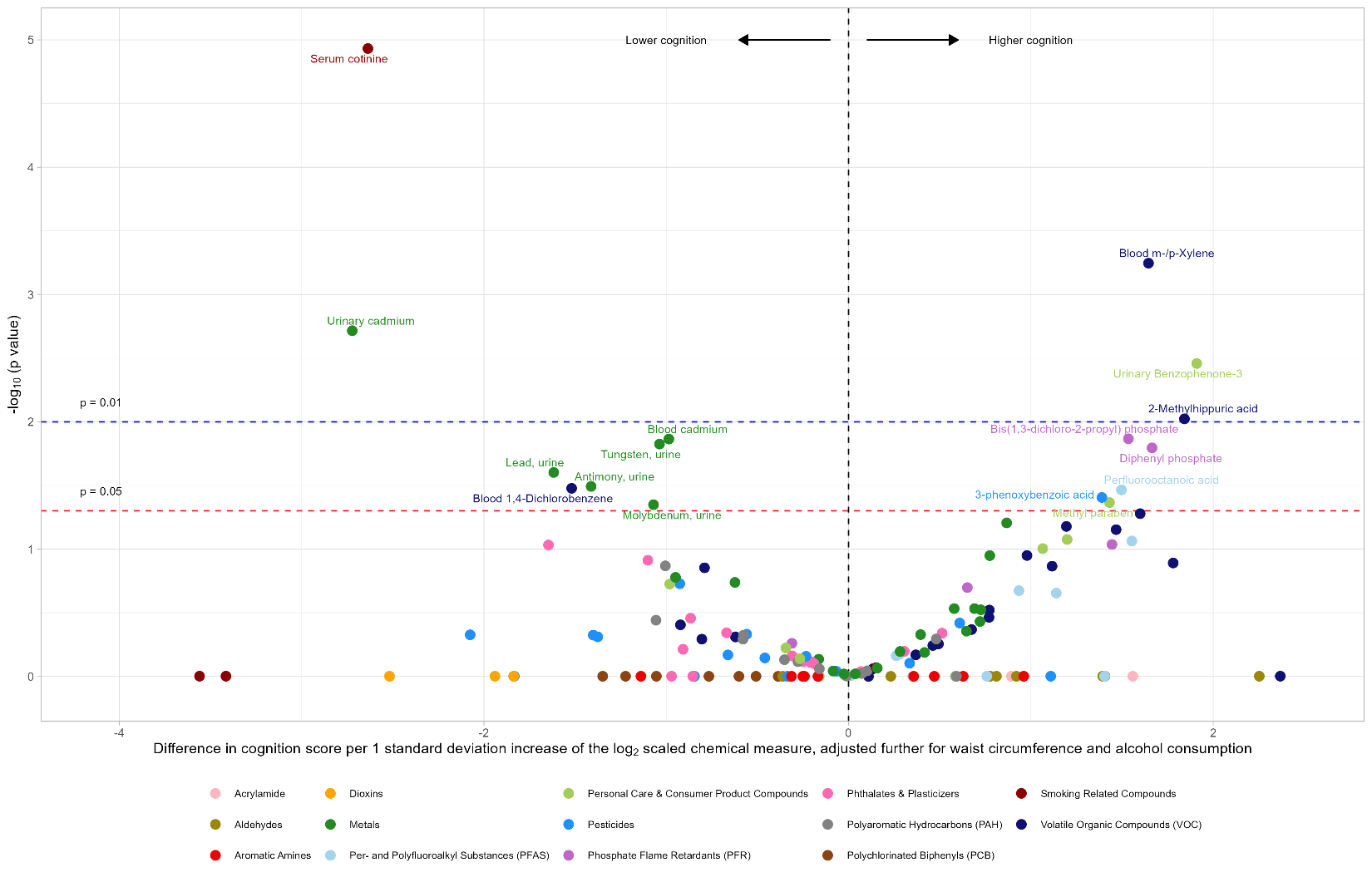

**Supplemental Table 7.** Additional covariate adjusted full results. Summary of survey-weighted linear regression models evaluating the association between chemical exposure biomarkers and Digit Symbol Substitution Test (DSST) scores, NHANES 1999-2000 and 2011-2014 (overall N = 4,982) for all chemicals.

Models were adjusted for age, sex, race/ethnicity, education, smoking status, blood cotinine, seafood consumption, NHANES cycle, urinary creatinine (urinary measures only), waist circumference, and alcohol consumption. Chemical exposure, blood cotinine, and urinary creatinine were log2 transformed and z-score standardized. P-values adjusted by controlling for the False Discovery Rate (FDR).

Available as a separate excel file: “Supplemental Table 7.xlsx”

**Supplemental Table 8.** Modified Poisson regression full results. Summary of survey-weighted modified Poisson regression models evaluating the association between chemical exposure biomarkers and mild cognitive impairment status in NHANES 1999-2000 and 2011-2014 (overall N = 4,982) for all chemicals.

Models were adjusted for age, sex, race/ethnicity, education, smoking status, blood cotinine, seafood consumption, NHANES cycle, and urinary creatinine (urinary measures only). Chemical exposure, blood cotinine, and urinary creatinine were log2 transformed and z-score standardized. P-values adjusted by controlling for the False Discovery Rate (FDR).

Available as a separate excel file: “Supplemental Table 8.xlsx”

**Supplemental Figure 5.** Volcano plot of associations between chemical exposures and mild cognitive impairment (MCI) among participants aged 60+ with estimated glomerular filtration rate ≥ 60 ml/min/1.73 m^2^ in the National Health and Nutrition Examination Survey (NHANES) cycles 1999-2000, 2011-2012, and 2013-2014. Relative risks (prevalence ratios) derived from modified Poisson regression models are plotted on the x-axis and represent a 1-standard deviation increase of the log_2_-transformed exposure measure. The black vertical line indicates a relative risk of 1, representing no association with MCI. P-values are plotted on the y-axis on a -log_10_ scale, so that p-values closer to 1 are at the bottom of the plot. The red horizontal line indicates the threshold of p = 0.05 and the blue horizontal line indicates the threshold of p = 0.01.

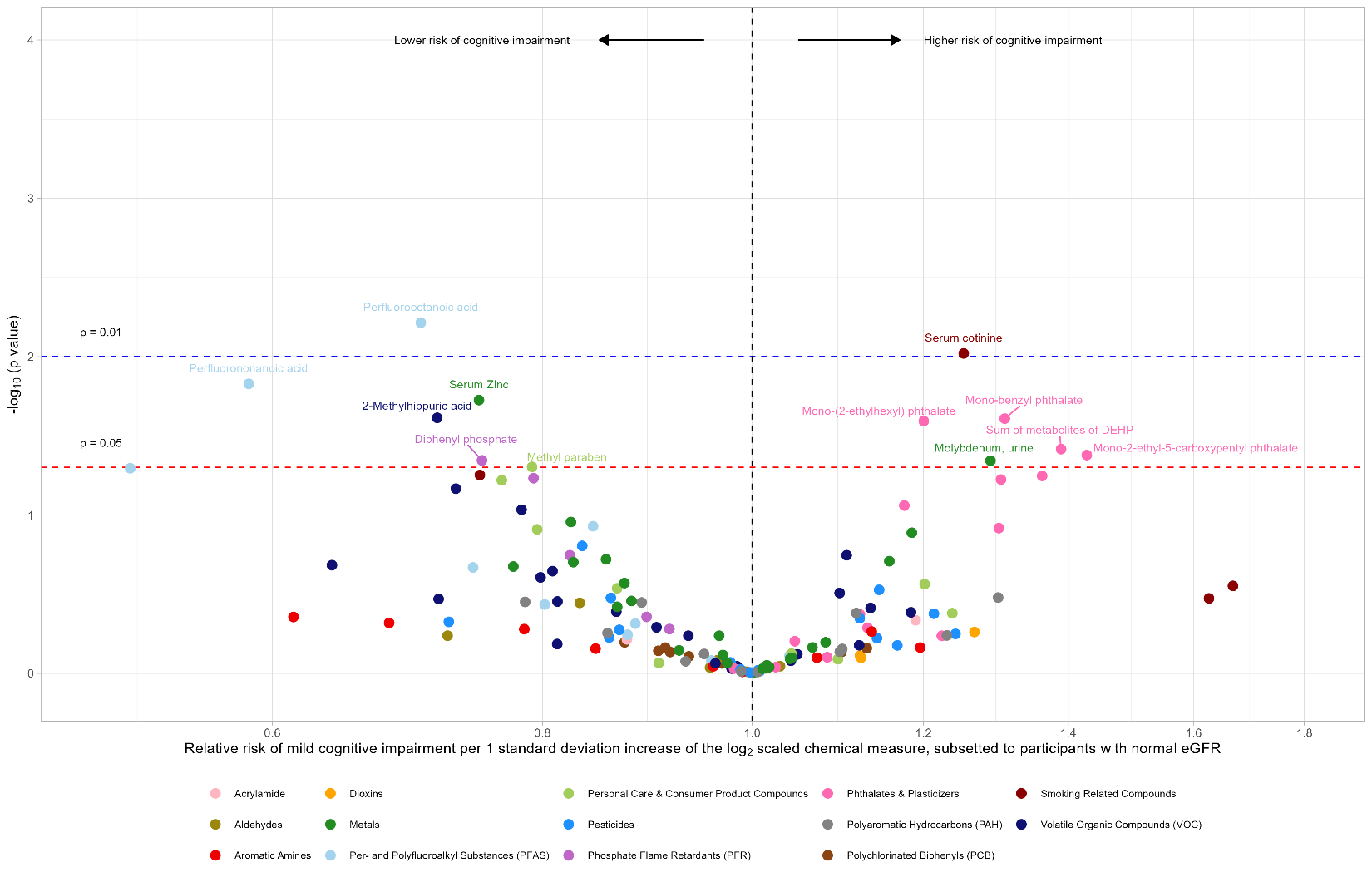

**Supplemental Table 9.** Modified Poisson regression among the subset of participants with normal estimated glomerular filtration rate (eGFR, ≥60 ml/min/1.73 m^2^) as a marker of kidney function. Summary of survey-weighted modified Poisson regression models evaluating the association between chemical exposure biomarkers and mild cognitive impairment status in NHANES 1999-2000 and 2011-2014 (normal eGFR N = 3,593) for all chemicals.

Models were adjusted for age, sex, race/ethnicity, education, smoking status, blood cotinine, seafood consumption, NHANES cycle, and urinary creatinine (urinary measures only). Chemical exposure, blood cotinine, and urinary creatinine were log2 transformed and z-score standardized. P-values adjusted by controlling for the False Discovery Rate (FDR).

Available as a separate excel file: “Supplemental Table 9.xlsx”
